# DETERMINATION OF THE ROUTINE SELF-CARE PRACTICES AMONG PATIENTS WITH DMTII IN KITUI COUNTY

**DOI:** 10.1101/2023.09.06.23295109

**Authors:** Mary Musembi, Catherine Syombua Mwenda, M Ramaligham Ramani

## Abstract

Worldwide, an estimated increase of 642 million patients will have been diagnosed with diabetes mellitus by the year 2035, out of this figure more than 90% will have type 2 diabetes mellitus. In Kenya the burden for the disease is equally on rise, consequently there is huge gap in knowledge deficient among the sick individuals and their families on diabetes management making most patients to present with poor self-care practices especially in rural settings where Resources are scarce. The study aims at developing a family-based intervention model to improve family participation in the management of patients with diabetes mellitus type II at Kitui County. A sample of 68 participants was required to participate in the study. Creating informative resources that highlight the importance of family involvement and provide guidance on DMTII self-care practices. A significant proportion of DMTII patients Out of the total sample size, 28 participants (representing 49.1% of the sample) reported that they monitor their blood sugars on a daily basis. On the other hand, 29 participants (50.9% of the sample) indicated that they do not engage in this practice. These monitoring overall health, reviewing medication plans, adjusting treatment as necessary, and receiving education on diabetes management. These findings emphasize the importance of targeted interventions and education to improve self-care practices among individuals with DMTII and address areas where adherence is lacking. While many individuals exhibit positive self-care behaviors, there are areas that require attention and improvement. Healthcare providers can utilize these findings to develop targeted interventions, patient education programs, and support systems to enhance self-care practices among DMTII patients. By promoting and facilitating optimal self-care activities, healthcare providers can help improve health outcomes and reduce the risk of complications for individuals living with DMTII.

## 1.0 Introduction

### 1.1 Background of the Study

Intervention done at the family level is the best self-care management strategy for people with the disease (ADA, 2017). Recent surveys have shown that most diabetes management occurs at home, where family members have a critical role in influencing care (WHO, 2012). Studies done by Orvik et al.. (2019) reported various reactions from family members upon learning that their kin has DMTII; for example, Families with little knowledge of disease management may feel distressed by the sickness of their kin; as a result, may not know what to do to support them, may not either understand the patient’s needs, may lack strategies to cope with the emotional aspect of the disease and therefore fail to participate in the care. American diabetes association,(2017). Evidence also shows that family participation in diabetes care can improve patients’ self-care management.

Consequently, recent studies have reported low levels of family participation in diabetes self-care management. (ADA, 2018), thus making family-based intervention the best strategy for disease management, as such interventions are designed to provide information, guidance, training, and support to not only the sick persons but also to family members who might be experiencing stress while dealing with patients with the disease (ADA, 2017). Like other countries in the developing world, Kenya continues to carry a heavy burden of this disorder, possibly reaching epidemic proportions(Word Health Organization, 2016). The crude prevalence of DMTII in the country has been documented to be 5.3 percent, with a higher number of cases at 16% in urban areas compared to 12% in rural parts. (El-busily et al.,2019). Interventions done at home, such as education, demonstrational empower not only the patients but also their family members to gain the necessary knowledge and skills, including communication, decision-making, and problem-solving, that enable them to actively participate in diabetes self-care practice at their homes and hence be able to reduce the fatal disease-related complications (Jones et al., 2018). Understanding factors that contribute to good diabetes self-care management can help guide in adapting to the best interventional strategies aimed at improving self-care practices, achieve better glycemic control and prevent diabetes-related complications(ADA, 2018)

### 2.0 Literature Review

### 2.1 The routine self-care Practices among patients with Diabetes Mellitus Type II

Self-care practices in patients with DMTII is significant in keeping the disease under control and prevents diabetes related complications(castro et al, 2018). World Health Organization (2018), defined self-care practice as the ability of an individual, family and the society to promote and maintain health, prevent occurrence of a disease and cope with the illness and related complications with or without any external support while self – management as a process where a patient uses the learnt abilities and skills to manage a chronic disease or risk factors. Recent demonstrations have enlisted that individuals with TIIDM have significantly low productivity and participatory rates

DMTII is a long term condition that requires an individual to make a variety of daily decisions on diabetes self-care management and perform significant self-care activities(WHO,2016). Self-care practices involve variety of areas including diet, exercise, drug, stress management, sleeping patterns, medical seeking behaviors (Tommky et al, 2018).In order for a patient to present with an effective self-care practices, good self-care management must be embraced (Brunisholz et al, (2016). Diabetes self-care behaviors are the practices embraced by persons with or at risk of diabetes mellitus aimed at managing their disease effectively on their own.(Tomky et al, 2016). While self-care is the care that includes any deliberate move aimed at looking at an individual’s physical, mental and emotional health however, self-management is characterized by patient’s decision and behaviors that they engage in any long term disorder that affects their well-being ((Bodenheimer, et al, 2016). Study done by (Nwanko, 2018) & Shrivastava et al, 2018), reported seven known types of diabetes self-care practices including consuming healthy diet, physical dynamics, self-glucose monitoring, adherence to proper medication, adoption of good problem solving skills, sound adapting capabilities and risk reduction modalities that an individual must perform routinely. Good diabetes self-care practices among the sick individuals can mysteriously alter the progression and prevent development of disease related complications. (Shrivastava et al, 2017). Individuals with DMTII have a significant responsibility of maintaining their own health through clear and concise communication with their families and occasionally seek for necessary advice as need arises (WHO, 2016).

### 3.0 Methodology

The study was based cross-sectional descriptive study design was employed. Mixed methods approach was used to collect both quantitative and qualitative data from study participants. The study was carried out in Kitui County. The county is located 170 km to the south East of Nairobi city, in former eastern province of Kenya. Its capital and largest town is Kitui with Mwingi serving as a major urban centre. Kitui county has a population of 1,136187 (2019 census). The study population comprised of all adult patients clinically diagnosed with diabetes mellitus Type II for a period of at least six months. A questionnaire is a research instrument that is used to collect data comprising of a number of questions for the study participants to respond on. A Structured interview questionnaire was used to collect qualitative data from focused group discussions while semi-structured questionnaire was used to collect quantitative data. The data was analyzed using SPSS version 25.

## 4.0 Results and Findings

### 4.1 Descriptive analysis on determination of the routine self-care practices among DMTII patients in Kitui county

Table 4 below indicated the analysis on the determination of the routine self-care practices among the DMTII patients and it was represented in form of frequencies and percentages and the results were tabulated as below;

**Table 1:**
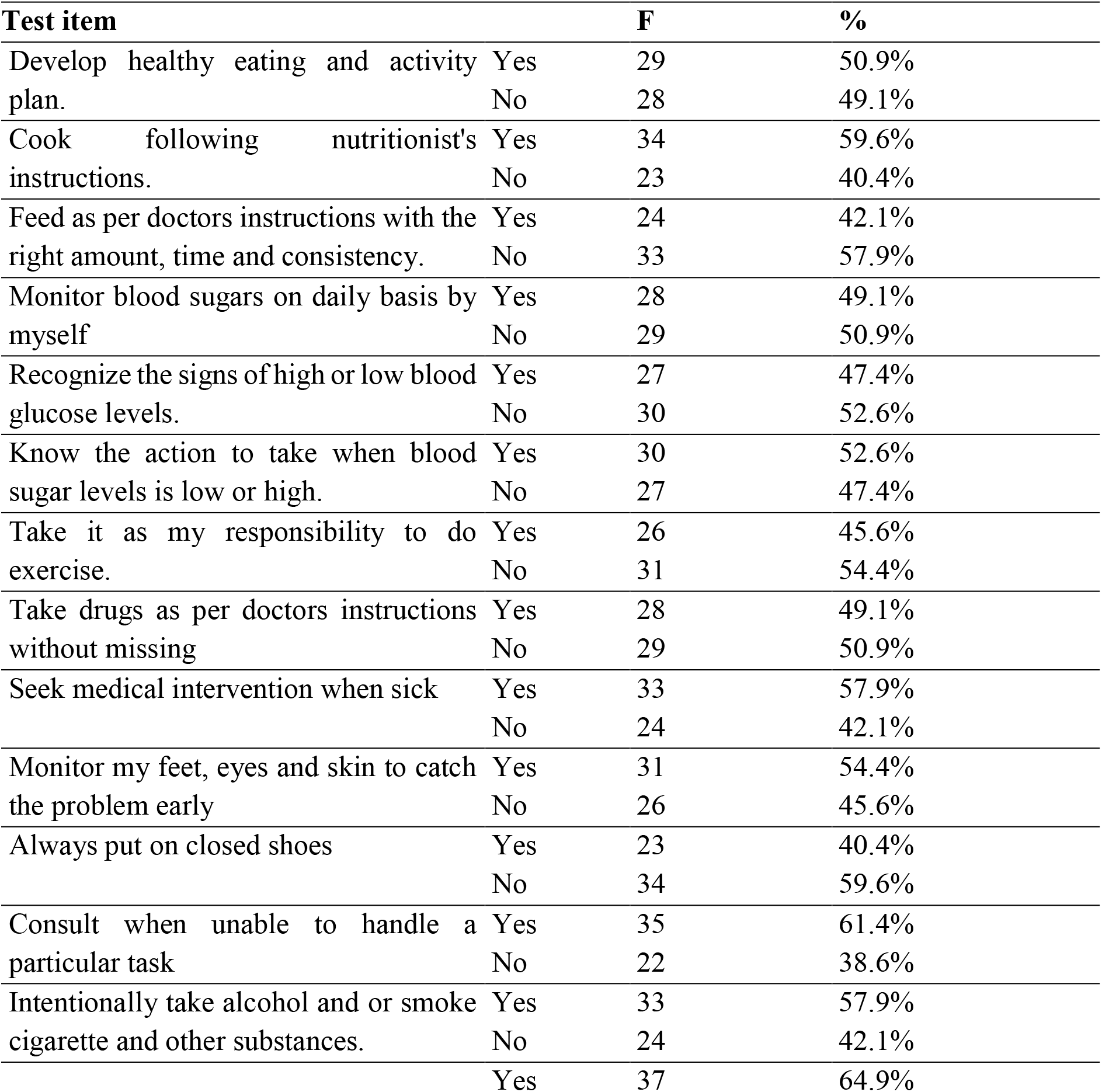
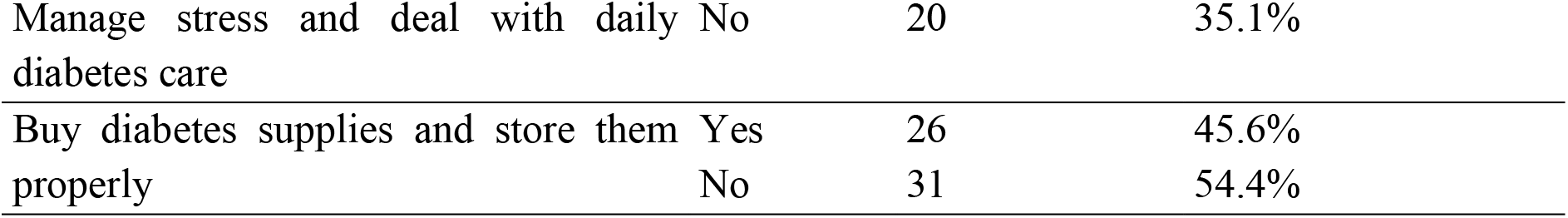
Descriptive analysis on determination of the routineself-care practices among DMTII patients in Kitui county.

### Source Field Data (2023)

Out of the total respondents, 29 individuals responded with “Yes,” indicating that they have developed a healthy eating and activity plan. This accounts for approximately 50.9% of the total respondents. On the other hand, 28 individuals responded with “No,” representing approximately 49.1% of the respondents who have not developed a healthy eating and activity plan. To determine the “Cook following nutritionist’s instructions,” two parameters were measured: “Yes” and “No.” The frequency of the “Yes” response was 34, which corresponds to a percentage of 59.6%. On the other hand, the frequency of the “No” response was 23, accounting for a percentage of 40.4%. These statistics provide insight into the participants’ adherence to the nutritionist’s instructions when cooking. The majority of participants (59.6%) reported following the instructions, while a significant portion (40.4%) indicated not following them. These findings suggest that a considerable proportion of individuals may have deviated from the prescribed instructions when preparing meals, indicating a potential area for further exploration or intervention to promote adherence to nutritional guidelines.

Among the respondents, 24 individuals (42.1%) answered “Yes,” indicating that they adhere to the nutritionist’s instructions for feeding. These individuals make conscious efforts to follow the prescribed guidelines provided by their doctors, ensuring that they provide the right amount of feed at the appropriate times, and maintain consistency in their feeding routine. On the other hand, 33 individuals (57.9%) responded with “No,” implying that they do not strictly follow the nutritionists’s instructions when it comes to feeding. These individuals may have various reasons for not adhering to the prescribed guidelines, such as personal preferences, challenges in maintaining consistency, or lack of awareness regarding the importance of following the instructions.

In the study, one of the test variables examined was the practice of monitoring blood sugars on a daily basis by oneself. The parameter used to measure this variable was a simple binary choice between “Yes” and “No.” Out of the total sample size, 28 participants (representing 49.1% of the sample) reported that they monitor their blood sugars on a daily basis. On the other hand, 29 participants (50.9% of the sample) indicated that they do not engage in this practice. The determine how respndents are able to detect or “Recognize the signs of high or low blood glucose levels” was administered to the participants, with two response options: “Yes” and “No.” Among the participants, 27 individuals responded “Yes,” indicating that they are able to detect/ recognize the signs of high or low blood glucose levels. This corresponds to a frequency of 27. The percentage of participants who responded “Yes” is calculated as 47.4%. On the other hand, 30 participants responded “No,” indicating that they were not able to recognize the signs of high or low blood glucose levels. 30 respondents answered “Yes,” indicating that they possess knowledge about the appropriate actions to take when blood sugar levels are low or high. This accounts for 52.6% of the total responses. On the other hand, 27 respondents answered “No,” indicating a lack of knowledge regarding the necessary actions. This represents 47.4% of the total responses. A total response of 26 individuals (45.6%) answered “Yes,” indicating that they perceive it as their responsibility to engage in exercise. On the other hand, 31 individuals (54.4%) answered “No,” indicating that they do not perceive it as their responsibility to exercise.

Out of the total number of respondents, 28 individuals (49.1%) answered “Yes,” indicating that they adhere to their doctor’s instructions and do not miss taking their prescribed drugs. On the other hand, 29 individuals (50.9%) answered “No,” indicating that they do not consistently follow their doctor’s instructions regarding medication and may occasionally miss taking their prescribed drugs. In examining the “Seek medical intervention when sick,” the parameters consist of “Yes” and “No.” Among the respondents, 33 individuals (57.9%) answered “Yes,” indicating that they actively pursue medical intervention when they are ill. Conversely, 24 individuals (42.1%) answered “No,” indicating that they do not seek medical intervention when they are sick. The study further indicated that 31 individuals (54.4%) answered “Yes,” indicating that they actively monitor their feet, eyes, and skin to catch any problems early. On the other hand, 26 individuals (45.6%) answered “No,” indicating that they do not engage in regular monitoring of their feet, eyes, and skin for early problem detection. Among the total number of respondents, 23 individuals (40.4%) answered “Yes,” indicating that they consistently wear closed shoes. On the other hand, 34 individuals (59.6%) answered “No,” indicating that they do not always put on closed shoes.

The study determined whether the patients were Consulting when unable to handle a particular task”this was evaluated based on the parameters “Yes” and “No.” Out of the total respondents, 35 individuals (61.4%) indicated that they consulted with others and relevant authorities when they were unable to handle a particular task. This implies that they are able to recognize the importance of seeking assistance or guidance when faced with challenges. Conversely, 22 individuals (38.6%) answered “No,” indicating that they prefer not to seek help and instead handle tasks independently. In analyzing the test variable “Intentionally take alcohol and/or smoke cigarettes and other substances,” the parameters include “Yes” and “No.” Out of the total number of respondents, 33 individuals (57.9%) answered “Yes,” indicating that they intentionally consume alcohol and/or smoke cigarettes and other substances. Conversely, 24 individuals (42.1%) answered “No,” indicating that they do not engage in intentional alcohol consumption or cigarette smoking.

Out of the total number of respondents, 37 individuals (64.9%) answered “Yes,” indicating that they are capable of managing stress and coping with the daily care required for diabetes. On the other hand, 20 individuals (35.1%) answered “No,” indicating that they struggle with managing stress and dealing with the demands of daily diabetes care. Out of the total number of respondents, 26 individuals (45.6%) responded with “Yes,” indicating that they do purchase diabetes supplies and ensure proper storage. Conversely, 31 individuals (54.4%) responded with “No,” indicating that they do not engage in buying diabetes supplies or storing them appropriately.

### 4.2 Inferential analysis on determination of the routine self-care practices among DMTII patients in Kitui county

Inferential analysis was determined using one sample t test among the patients who were found to have DMTII in Kitui county on the self-care practices and the results were presented on table 5 below;

**Table 2:**
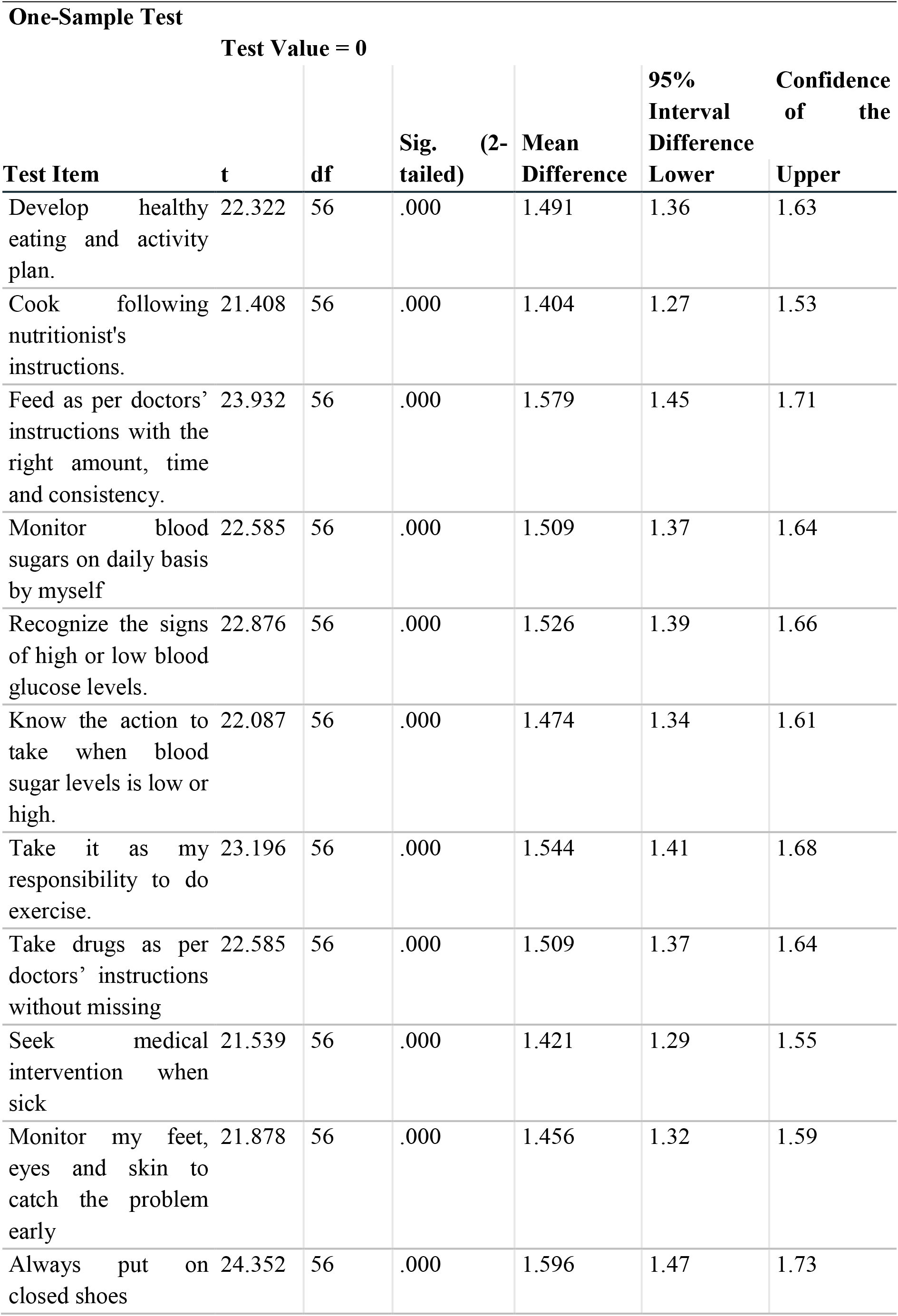
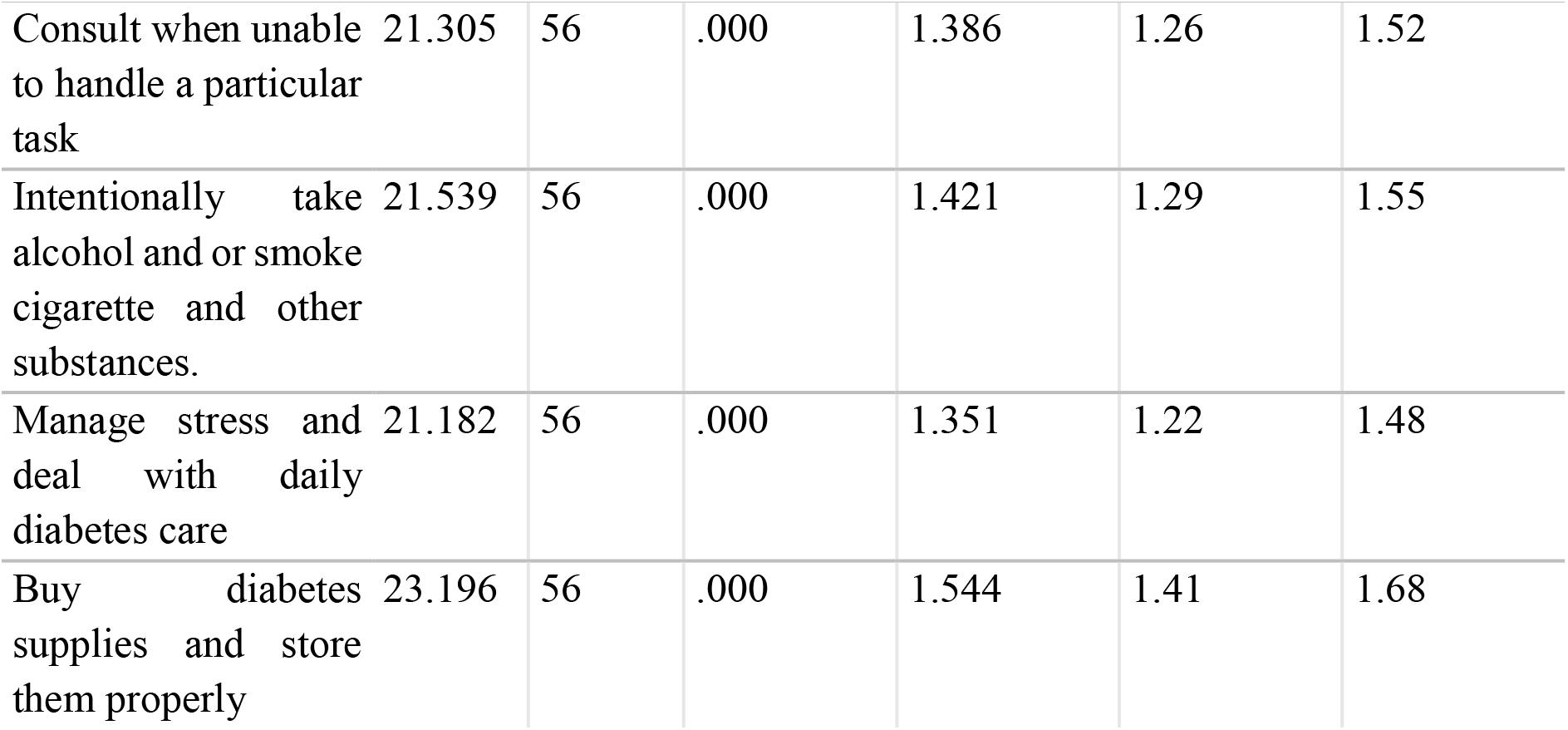
One sample t-test self-care practices among DMTII patients One-Sample Test.

### Source Field Data (2023)

To determine the “Develop healthy eating and activity plan” was analyzed using a one-sample test, which produced the following results: a t-value of 22.322, a significance level (Sig.) of .000 this indicated that that the obtained results were statistically significant and a mean difference of 1.491 this indicated that the extent of the change or effect related to the development of healthy eating and activity plans. The t-value indicates the magnitude of the difference between the observed mean and a hypothetical mean, with higher values indicating a larger difference.

The study determined that “Always put on closed shoes” was analyzed using a one-sample test, which yielded a t-value of 24.352. This indicated the t-value is significantly different from zero, with a p-value (Sig. 2-tailed) of .000, indicating a highly significant result. The mean difference between the sample mean and the hypothesized population mean is 1.596. Since the t value was highest the study concluded there was a statistical significant difference between the observed sample mean and the hypothesized population mean, supporting the notion that individuals tend to consistently put on closed shoes.

The “Manage stress and deal with daily diabetes care” was analyzed using a One-Sample Test and had the least t value according to table 5 above. The t-value obtained was 21.182, indicating a significant difference between the observed mean and the hypothesized population mean. The significance level (Sig. 2-tailed) was recorded as .000, which is below the conventional threshold of .05, suggesting strong evidence against the null hypothesis. The mean difference of 1.351 signifies the extent of deviation between the observed sample mean and the hypothesized population mean. Overall, these results indicate that there is a substantial difference in how individuals manage stress and handle their daily diabetes care, as supported by the significant t-value and low p-value.

The study concluded that since all the parameters used to determine self-care practices among DMTII patients had a statistical relationship since all the p value obtained were 0.000 which were less than 0.05 hence indicated statistical relationship between self-care practices and contacting DMTII among patients in Kitui County.

### 4.3 Thematic analysis on determination of the routine self-care practices among DMTII patients in Kitui county

The study determined the self-care practices among DMTII patients and the following are some of the factors that were indicated by the respondents that would help them to camber with self-care practices;

> *“Assessing the patient’s understanding of their condition and knowledge about diabetes management. Evaluate their awareness of the importance of self-care practices, including diet, physical activity, medication adherence, and blood glucose monitoring*.*”*
>
> *“Assess the patient’s adherence to prescribed medications, including oral hypoglycemic agents or insulin. Discuss any challenges they may face in taking medications regularly, such as forgetfulness, side effects, or cost-related concerns*.*”*
>
> *“Assess the patient’s emotional well-being and their coping mechanisms. Diabetes can be emotionally challenging, so it is crucial to address any feelings of depression, anxiety, or diabetes-related distress. Encourage support systems, counseling, or appropriate referrals if needed*.*”*
>
> *“Exploring the patient’s stress levels and their ability to manage stress effectively. Discuss stress management techniques such as relaxation exercises, meditation, counseling, or support groups that can help in coping with the emotional and psychological impact of diabetes*.*”*
>
> *“Assessing the patient’s adherence to prescribed medications, including oral hypoglycemic agents or insulin. Discuss any challenges they may face in taking medications regularly, such as forgetfulness, side effects, or cost-related concerns*.*”*
>
> *“Evaluating the patient’s current level of physical activity and their willingness to engage in regular exercise. Discuss the benefits of exercise in managing diabetes, including improved insulin sensitivity, weight management, cardiovascular health, and stress reduction*.*”*
>
> *“Evaluating the patient’s social network and support systems. Identify the availability of family, friends, or support groups that can offer encouragement, assistance, and motivation in their self-care efforts*.*”*

The study indicated that it is essential to approach the determination of self-care practices among DMTII patients holistically, considering these aspects and collaborating with the patient to develop an individualized self-care plan that suits their unique needs and circumstances.

## 5.0 Discussion, Conclusion and Recommendation

### 5.1 Discussion

The findings of this study indicate that a substantial number of DMTII patients adhere to recommended self-care practices, such as medication adherence, blood glucose monitoring, and regular healthcare check-ups. However, areas of improvement are identified, including dietary habits, physical activity, and consistent healthcare visitation. Strategies to enhance self-care practices should focus on education, patient empowerment, behavioral change techniques, and tailored interventions addressing specific challenges faced by DMTII patients.

A significant proportion of DMTII patients Out of the total sample size, 28 participants (representing 49.1% of the sample) reported that they monitor their blood sugars on a daily basis. On the other hand, 29 participants (50.9% of the sample) indicated that they do not engage in this practice. These monitoring overall health, reviewing medication plans, adjusting treatment as necessary, and receiving education on diabetes management. Consulting when unable to handle a task was reported by 61.4%, while 38.6% preferred not to seek help. Intentional alcohol consumption and smoking were reported by 57.9%, while 42.1% did not engage in these behaviors. Managing stress and daily care for diabetes was reported by 64.9%, while 35.1% struggled with these aspects. Purchasing diabetes supplies and proper storage was reported by 45.6%, while 54.4% did not engage in these practices. These findings emphasize the importance of targeted interventions and education to improve self-care practices among individuals with DMTII and address areas where adherence is lacking.

## 5.2 Conclusion

This study provides valuable insights into the routine self-care practices among DMTII patients. While many individuals exhibit positive self-care behaviors, there are areas that require attention and improvement. Healthcare providers can utilize these findings to develop targeted interventions, patient education programs, and support systems to enhance self-care practices among DMTII patients. By promoting and facilitating optimal self-care activities, healthcare providers can help improve health outcomes and reduce the risk of complications for individuals living with DMTII.

### 5.3 Recommendations

Develop interventions to address barriers to regular healthcare check-ups and promote consistent engagement with healthcare providers.

Employ technology-based solutions, such as mobile applications or telehealth services, to enhance self-care practices and facilitate remote monitoring and support.

## Data Availability

Field data collected from Kitui County, Kenya

https://drive.google.com/file/d/1dwCVNwmuN3PfPeCv1lf17X6F9tXQxjG6/view?usp=sharing

